# Assessing Etiologic Heterogeneity for Multinomial Outcome with Two-Phase Outcome-Dependent Sampling Design

**DOI:** 10.1101/2022.07.20.22277805

**Authors:** Sarah A. Reifeis, Michael G. Hudgens, Melissa A. Troester, Michael I. Love

## Abstract

Etiologic heterogeneity occurs when distinct sets of events or exposures give rise to different subtypes of disease. Inference about subtype-specific exposure effects from two-phase outcome-dependent sampling data requires adjustment for both confounding and the sampling design. Common approaches to inference for these effects do not necessarily appropriately adjust for these sources of bias, or allow for formal comparisons of effects across different subtypes. Herein, using inverse probability weighting (IPW) to fit a multinomial model is shown to yield valid inference with this sampling design for subtype-specific exposure effects and contrasts thereof. The IPW approach is compared to common regression-based methods for assessing exposure effect heterogeneity using simulations. The methods are applied to estimate subtype-specific effects of various exposures on breast cancer risk in the Carolina Breast Cancer Study.

## 1 Introduction

Two-phase outcome-dependent sampling (ODS) designs are often utilized in observational studies as a cost effective way to estimate exposure effects on an outcome of interest. Two-phase sampling designs entail two steps. First, a random sample of individuals is drawn from a population; outcome and covariate data are collected on these individuals. Second, individuals are stratified according to the first-phase data, and simple random samples are drawn from each stratum, with known stratum-specific selection probabilities. Additional covariates, possibly including exposures of interest, are collected on the individuals selected for this second phase (Neyman, 1938). Assessing exposure effects in two-phase observational studies is challenging because adjustment is needed for both confounding and unequal probability sampling.

ODS is often performed with respect to a binary outcome, such as in case-control studies. However, many disease outcomes are dichotomized for ease of analysis but are not truly binary. For example, many subtypes of disease exist for particular cancers and it may be of interest to researchers to compare exposure effects for these different subtypes. The effect of a given exposure on cancer risk overall may be null, but its effect on certain subtypes could be non-null, as it could confer an increase in risk for one or more subtypes, while a decrease for others. Etiologic heterogeneity refers to the situation where different sets of events or exposures may give rise to the different subtypes of disease, or more subtly that the degree of risk conferred may differ across subtypes of disease. Such heterogeneity is of particular interest in cancer epidemiology (Zabor and Begg, 2017). The ability to determine whether or not a given exposure raises cancer risk for a particular subtype more than the other subtypes can allow for identification of previously unknown relationships, and can help to generate new mechanistic hypotheses. For example, when assessing environmental or behavioral risk factors for breast cancer, investigators might want to determine: Is there a difference in the effect of oral contraceptive use on risk of developing estrogen receptor-positive compared to estrogen receptor-negative breast cancer? If so, how big is the difference? Answers to these kinds of questions can be informed by point and interval estimates of subtype-specific exposure effects as well as hypothesis tests for equality of these effects.

When considering risk factor effects on subtypes of disease, it is important to distinguish between genetic and environmental/behavioral risk factors. Analyses of genome-wide association studies often rely on the assumption that individuals in the study are not related, or that mixed models sufficiently capture relatedness due to ancestry differences in the population under study (Sul et al., 2018). Having adjusted for population structure, it is sometimes assumed that genetic risk factors are not influenced by other variables. For example, Zhang et al. (2020), Zhang et al. (2021), and Ahearn et al. (2022) have recently studied etiologic heterogeneity with genetic risk factors using mixed-effect models that allow for, but do not require, inclusion of additional covariates in the model for adjustment.

On the other hand, environmental or behavioral risk factors and exposures are often influenced by many other variables. As a result, confounding can generally be expected when estimating environmental and behavioral exposure effects on disease subtypes from observational data. Zabor and Begg (2017) compared multinomial logistic regression with methods from Chatterjee (2004), Rosner et al. (2013), and Wang et al. (2015) that were developed for investigation of etiologic heterogeneity, but this comparison did not evaluate these methods for confounding adjustment. Benefield et al. (2019) estimated various environmental exposure effects on risk of developing subtypes of breast cancer using regression and including confounders as predictors in the models. While the aforementioned methods allow for incorporating confounders as predictors in a regression model, they each rely on the strong assumption that the confounders and exposure do not interact to influence the outcome.

Inverse probability weighting (IPW) has been proposed to adjust for both confounding and biased sampling when estimating exposure effects for studies with ODS (Wang et al., 2009), but not in the context of a multinomial outcome. Additionally, IPW has been used for estimation of exposure effects on multinomial outcomes (Richardson et al., 2018), but not in the context of ODS. When estimating exposure effects on multinomial outcomes for studies using ODS, methods are needed that (i) appropriately adjust for confounding, (ii) account for the sampling design, and (iii) accommodate the multinomial nature of the outcome to answer questions of exposure effect heterogeneity.

In this paper methods are considered which address (i) – (iii). In particular, in Section 2 an IPW approach is developed that extends a method from Wang et al. (2009) to ODS studies where the outcome is multinomial for inference about subtype-specific exposure effects, and two common regression-based approaches are also described. These methods are compared in Section 3 using simulation studies and data from the Carolina Breast Cancer Study (CBCS). Section 4 concludes with a review and discussion of the main results, and addresses limitations and future directions for this research. Appendix A of the Supplementary Materials provides further details of the methods introduced in Section 2. Additional simulation study results are included Appendix B, and Appendix C contains SAS code for analyzing an example data set with each method considered.

## 2 Methods

Consider a two-phase ODS design where the goal is to estimate the effects of a binary exposure *A* on a multinomial outcome *Y*, where *Y* takes values *k* = 0, …, *K* for *K* ≥ 1. The outcome levels *k* are referred to as (disease) subtypes, where *k* = 0 denotes no disease. Let *L* = (*L*_1_, *L*_2_) where *L*_1_ and *L*_2_ denote vectors of covariates observed for all first-and second-phase individuals, respectively. Define *Y*_*k*_ = *I*(*Y* = *k*) to be the indicator of experiencing subtype *k* ∈ {0,…, *K*}. Note that 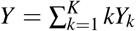. Let *S* be the indicator of selection for the second phase, and *g*(*Y, L*_1_, *A*) = *P*(*S* = 1|*Y, L*_1_, *A*) represent the known second-phase selection probabilities. Suppose we observe *m* i.i.d. copies of *O*_*i*_ = (*L*_1*i*_, *S*_*i*_*L*_2*i*_,*Y*_*i*_, *A*_*i*_, *S*_*i*_) for *i* = 1, …, *m*. Assume *S* ⊥ *L*_2_ | *Y, L*_1_, *A*, i.e., conditioning on the first-phase data, selection is independent of the covariates observed in the second phase of sampling. Let 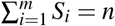 denote the number of individuals selected in the second phase of sampling.

Exposure (or risk factor) effects can be defined using potential outcomes (or counterfactuals). Let *Y*^*a*^ and 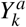be the potential outcome and potential subtype indicator if, possibly counter to fact, an individual had exposure *a* for *a* = 0, 1. Let 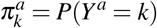represent the counterfactual risk of developing subtype *k* = 1, …, *K* had the exposure and let been *a* and let 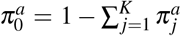 for *a* = 0,1. Let *e*_a_ (*L*) = *P*(*A* = *a*|*L*) represent the probability of exposure *a* given covariate vector *L*, for *a* = 0, 1. Ultimately our goal is to draw inference about the effects of *A* on the risk of developing different disease subtypes, which may be defined in terms of contrasts in the subtype-specific counterfactual risks 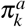.

Assume 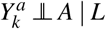 for *a* = 0, 1 and *k* = 1, …, *K* (conditional exchangeability), 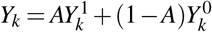 for *k* = 1, …, *K* (causal consistency), and *e*_*a*_(*l*) *>* 0 for *a* = 0, 1, and all *l* such that *dF*_*L*_(*l*) *>* 0 (positivity), where in general *F*_*X*_ denotes the cumulative distribution function (CDF) of *X*. Also assume that every individual has a nonzero second-phase sampling probability, i.e., *g*(*y, l*_1_, *a*) *>* 0 for all *y, l*_1_, and *a* where 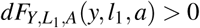. In combination with the other aforementioned assumptions, knowing these selection probabilities permits identifiability of the risks in the original population for the *K* non-reference subtypes, so functions of the risk (e.g., the risk difference, risk ratio, and relative risk ratio) are also identifiable.

There are various possible estimands of interest to quantify etiologic heterogeneity. Some estimands are *covariate-conditional*, meaning that the estimands quantify the exposure effects within strata of individuals with the same values of the covariates *L*. Estimands which are not covariate-conditional are referred to as *marginal* estimands. When the research question of interest is best answered by estimating a single quantity describing the population of interest, marginal estimands may be preferred. All estimands considered here are *subtype-conditional*, meaning that the estimand for each subtype *k* is conditional on the outcome being subtype *k* or no disease (i.e., subtype 0). This quality is desirable because it allows the “no disease” subtype to serve as a common reference outcome, facilitating comparisons between subtypes of disease. In Section 2.1 we consider IPW of a marginal structural model (MSM) where the estimands are marginal, whereas in Section 2.2 we consider regression-based estimators where the corresponding estimands are covariate-conditional.

### 2.1 Inverse Probability Weighting

Consider the following multinomial MSM

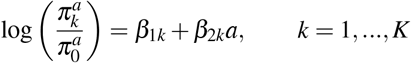

or equivalently 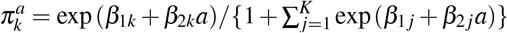. The parameter *β*_2*k*_ can be interpreted as a log relative risk ratio (RRR) or log subtype-conditional odds ratio (OR) by noting that

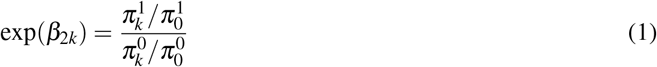

The numerator of (1) is the risk ratio comparing the risk of subtype *k* versus the risk of no disease had everyone been exposed; the denominator of (1) has a similar interpretation had everyone been unexposed. Thus exp(*β*_2*k*_) is the causal RRR of developing subtype *k* relative to no disease had everyone been exposed versus unexposed.

The RRR being less than one indicates exposure is protective against developing subtype *k* relative to no disease. Conversely if the RRR is greater than one, exposure confers increased risk of developing subtype *k* relative to no disease. Additionally, exp(*β*_2*k*_) can be interpreted as the causal subtype-conditional OR because the risk ratios in the numerator and denominator of (1) can be equivalently expressed as conditional odds, i.e., 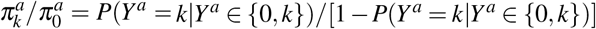 for *a* ∈ {0, 1} and *k* ∈ {1,…, *K*}. Thus the numerator of (1) is the odds of developing subtype *k* had everyone been exposed, among those who would develop subtype *k* or no disease, and the denominator has an analogous interpretation.

The parameters of the MSM can be estimated by weighted maximum likelihood as follows. First estimate *e*_*a*_(*L*) by 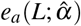using weighted maximum likelihood, where 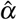represents estimated coefficients from the propensity score model, as described in Appendix A.1 of the Supplementary Materials. Then for each individual compute the weight 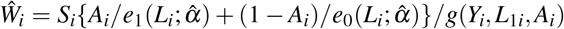. Next fit the following multinomial logistic regression model

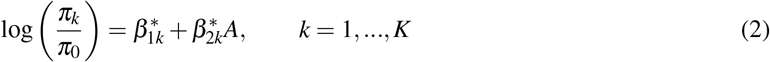

using weighted maximum likelihood with weights 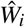 where *π*_*k*_ = *P*(*Y* = *k*|*S* = 1, *A*) is the probability of developing subtype *k* given second-phase selection and *A*, for *k* = 1, …, *K*, and 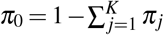. Let 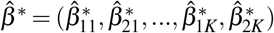 denote the weighted maximum likelihood estimator (MLE) of the parameters in model (2). The estimator 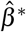 is consistent for *β* = (*β*_11_, *β*_21_, …, *β*_1*K*_, *β*_2*K*_) and asymptotically normal; see the proof in Appendix A.2 of the Supplementary Materials.

There are several methods available in standard software to estimate the covariance matrix of the estimated coefficient 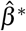. One approach to covariance matrix estimation entails a Taylor series (TS) approximation (Binder, 1983), which appropriately accounts for biased sampling and is the default method used in the *SURVEYLOGISTIC* procedure in SAS (SAS Institute Inc., 2017). In the setting described here, the TS estimator is consistent for the asymptotic covariance matrix when the propensity score is known and is proportional to the commonly used Huber-White “robust” sandwich variance estimator; see Appendix A.3 of the Supplementary Materials for details. A second approach to covariance matrix estimation is the nonparametric bootstrap (Mashreghi et al., 2016), which is also available in the *SURVEYLOGISTIC* procedure. For the bootstrap estimator in this context, random samples of size *n* are drawn with replacement from {*O*_*i*_ : *S*_*i*_ = 1, *i* = 1, …, *m* }, the weighted MLE 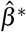 is computed for each replicate sample, and the covariance is estimated by the empirical covariance of 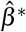 across the replicate samples. Note for this bootstrap procedure that 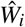 is estimated once for the original sample, and is not re-estimated for each replicate sample. While commonly used in the context of survey sampling, neither this bootstrap procedure nor the TS estimator account for estimation of the IPW weights. Alternatively, a nonparametric bootstrap estimator accounting for IPW weight estimation can be used, where the resampling process proceeds as described for the first bootstrap estimator but the propensity score is estimated for each replicate sample prior to computing 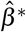. Variance estimates of the estimated subtype-conditional ORs can then be used to construct Wald confidence intervals (CIs).

Finally, it may be of interest to construct hypothesis tests comparing subtype-conditional ORs (or equivalently RRRs). *F*-tests may be used for formal comparisons of subtype-conditional ORs. For example, the test of no exposure effect heterogeneity across subtypes of disease, i.e., *H*_0_ : *β*_21_ = … = *β*_2*K*_, may be of interest. The test statistic 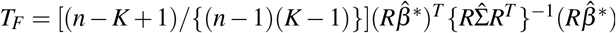may be used to test the null hypothesis *H*_0_, where *R* is a contrast matrix with *K* − 1 rows representing the structure of *H*_0_ and 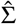 is an estimated covariance matrix (e.g., estimated using the TS method) evaluated at 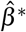. If the estimator 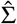 is consistent, then in large samples *T*_*F*_ will have approximate distribution *F*(*K* 1, *n K* + 1) under the null hypothesis. Similarly, the researcher may wish to evaluate exposure effect heterogeneity for subtypes *j* and *k* by modifying *R* and using the *F*-test of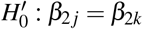.

### 2.2 Regression with Sampling Weights or Offsets

An alternative to IPW commonly used to estimate the effects of *A* on different subtypes entails using regression as described below. Let *X* = (1, *A, L*_1_, *L*_2_) and consider the following multinomial regression model

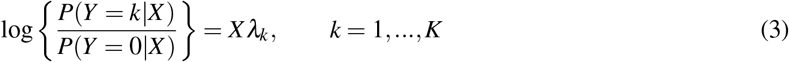

where 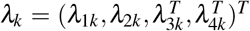is a column vector. Assuming conditional exchangeability and that the model (3) is correctly specified, exp(*λ*_2*k*_) represents the subtype-conditional OR of developing subtype *k* had everyone been exposed versus unexposed, within strata defined by *L*. Note that there are no interaction terms between the exposure *A* and the covariates *L* in model (3), such that the subtype-conditional OR is constant across strata. Since the OR is not generally collapsible (Greenland et al., 1999) over levels of the covariates *L*, the parameter *λ*_2*k*_ need not equal the parameter *β*_2*k*_ of the marginal structural model (2).

The parameters of model (3) can be estimated using either selection weights or an offset term to account for ODS. The selection weight for an individual *i* is defined as the inverse of their sampling probability, i.e., 1*/g*(*Y*_*i*_, *L*_1*i*_, *A*_*i*_), which is known by design; see Appendix A.4 of the Supplementary Materials for details of fitting the regression model with sampling weights. The offset approach (Weinberg and Wacholder, 1990) is applicable for study designs where the log relative risk of selection for the second phase is linear in *X*, i.e.,

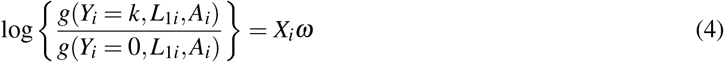

for some vector of constants *ω*. Because the sampling probabilities are known, *ω* will be known and therefore *X*_*i*_*ω* can be evaluated for each individual and used as an offset term in the regression model; see Appendix A.5 of the Supplementary Materials for details. Note the offset term does not depend on the value of *k* because here it is assumed that selection is performed with respect to the dichotomized outcome where 0 is the reference category and the selection probabilities for all subtypes *k* = 1, …, *K* are equal, i.e., *S* ⊥ {*Y, L*_2_} | *I*(*Y >* 0), *A, L*_1_.

Using either selection weights or the offset term (4), the parameters of model (3) can be estimated by fitting the Model

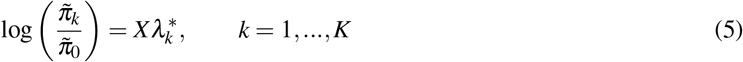

where 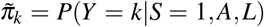is the probability of developing subtype *k* among the selected given *A* and *L* for 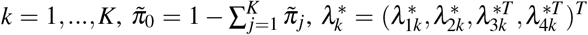is a column vector, and 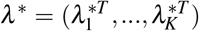Let 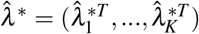represent the MLE of *λ* ^∗^ when fitting model (5) with sampling weights, and 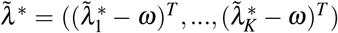denote the MLE of *λ* ^∗^ when fitting model (5) with the offset term. Then 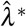 and 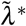are consistent for 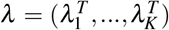and asymptotically normal; see Appendices A.4 and A.5 of the Supplementary

Materials for details. As with IPW, covariance matrix estimation for the regression approaches can be carried out with the TS or non-parametric bootstrap methods. Wald CIs for the estimated subtype-conditional ORs can be constructed using the variance estimates computed by these methods. Similarly, *F*-tests of any null hypothesis of interest that can be expressed as a contrast of model parameters may be performed. Note the null hypotheses constructed with covariate-conditional parameters *λ* differ from the null hypotheses described in Section 2.1, which are constructed using marginal parameters *β*.

### 2.3 Simulation Studies

Simulation studies were conducted to compare the finite-sample performance of the IPW and regression methods described above. Four phase one datasets were simulated, with the target parameters of interest defined specifically with respect to the *m* individuals in the first phase; this simulation design mimics the breast cancer study described in the following section. First-phase data sets of size *m* = 20, 000 and *m* = 200, 000 were simulated for each of two simulation study scenarios. The first-phase covariate *L*_1_ was generated from Bern(0.4), i.e., Bernoulli with expected value 0.4, and the second-phase covariate *L*_2_ from a Poisson distribution with expected value 3. The exposure *A* was generated unconditionally for the scenario with no confounding from Bern{expit(1)}, and conditional on *L*_1_ = *l*_1_, *L*_2_ = *l*_2_ for the scenario with confounding from Bern{expit(−1+ *l*_1_ +0.5*l*_2_)}. The potential outcomes *Y*^*a*^, *a* = 0, 1 were multinomial with three levels {0, 1, 2}. For the no confounding scenario, *Y*^*a*^ was generated unconditionally with 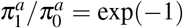 and 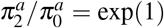. For the confounding scenario, *Y*^*a*^ was generated conditional on *L* = (*L*_1_ *L*_2_ with *P*(*Y*^*a*^ = *y*|*L*_1_ *L* = *l*) = *p* where *p /p* = exp(−1 + 1.5*a* −0.5*l*_1_ + 0.75*l*_2_ 0.75*al*_1_ 0.5*al*_2_), *p*_2_*/p*_0_ = exp(1.5 −0.5*a* + 0.75*l*_1_− 0.25*l*_2_ + 0.25*al*_1_ − *al*_2_), and *p*_0_ = 1 *p*_1_ *p*_2_. Five hundred second-phase samples of size *n* = 2, 000 were drawn from each of the first-phase data sets, resulting in sampling percentages of 10% and 1% for the first-phase data sets. The sampling weights were dependent on *L*_1_ and the dichotomized observed outcome *I*(*Y >* 0), but not the exposure *A*, and were defined such that approximately half of the observations within each second-phase data set had *Y* = 0; see Section C.1 of the Supplementary Materials for further details. Subtype-conditional ORs and their corresponding confidence intervals were estimated for subtypes 1 and 2, using each of the methods described in Section 2. All analyses here and in the following section were conducted with SAS software, Version 9.4 using the *SURVEYLOGISTIC* procedure (SAS Institute Inc., 2017) and the *%BOOT* macro.

Choosing the marginal subtype-conditional OR as the target parameter, the empirical bias of the estimated ORs and 95% CI coverage and width were computed, and the results were aggregated over the 500 simulated second-phase data sets. Each OR estimator’s variance was estimated using Taylor Series and bootstrap estimators. For the IPW estimator, the covariance matrix was estimated using the bootstrap estimator with and without accounting for estimation of the propensity score. All bootstrap covariance matrix estimates were computed using 250 replicates.

### 2.4 CBCS Analysis

The methods described above were applied to the CBCS Phase I-II data for estimation of subtype-conditional effects of several exposures on breast cancer. These data have been described and analyzed previously (Newman et al., 1995; Benefield et al., 2019). Phase I (1993-1996) and II (1996-2001) of CBCS were population-based case-control studies, where “Phase” here refers to the distinct time periods over which the CBCS was conducted and does not indicate phase with respect to two-phase ODS. Individuals with breast cancer (“cases”) were recruited into CBCS from the North Carolina Central Cancer Registry, and “controls” that represent the source population from which the cases were selected were recruited from Department of Motor Vehicles (DMV) and Medicare records. Age, race, and breast cancer status were known for all individuals in these records. Breast cancer subtype, exposures, and other covariate information were not recorded in the cancer registry, DMV, or Medicare records, and thus were only obtained for those individuals selected into the CBCS. Individuals in the cancer registry or in the DMV or Medicare databases were assumed to constitute a random sample of the population of interest, namely women from central North Carolina. Thus these individuals constitute the first-phase sample and participants in the CBCS comprise the second-phase sample in this two-phase ODS study.

Breast cancer subtypes were defined as in Benefield et al. (2019), namely based on estrogen receptor (ER) status (positive or negative) and tumor suppressor gene p53 status (positive or negative) obtained from genomic analysis of the breast tumor sample. The dichotomized exposures considered were BMI (≥ 25 / *<* 25), breastfeeding (ever/never), oral contraceptive (OC) use (ever/never), and parity (parous/nulliparous). The selection probabilities were fixed by study design and were dependent on study Phase (I or II), age, race, and breast cancer status. Note that in Section 2 these probabilities were allowed to depend on the exposure *A*, but for the CBCS data they are not dependent on any of the exposures considered. For all exposures considered, the set of variables assumed sufficient for confounding adjustment (i.e., conditional exchangeability) included age, race, and age at menarche. For the parity and breastfeeding exposures, the adjustment set also included BMI and OC use. For the BMI exposure, the adjustment set also included OC use. Finally, when OC use was the exposure, parity was also included in the adjustment set. Complete data on 2,789 individuals (pooled over Phases I and II) were analyzed.

## 3 Results

### 3.1 Simulation Studies

Results from the simulation studies described in Section 2.3 are presented in Table 1 for the setting where 1% of observations were selected from the first phase; results for 10% sampling are presented in Appendix B of the Supplementary Materials. In the first simulation scenario there was no confounding, and the bias, coverage, and CI widths were comparable across all methods as expected. In the second simulation scenario there was confounding as well as interaction between the exposure *A* and the covariates *L* in the data generating outcome regression model. The regression methods exhibited substantial bias and poor coverage for the marginal subtype-conditional OR for subtype 1 (*OR*_1_) but not for subtype 2 (*OR*_2_). On the other hand, the IPW estimator was approximately unbiased and the corresponding CIs achieved nominal coverage levels. These results accord with expectation in that the regression estimators are not necessarily expected to perform well when drawing inference about marginal effects in the presence of confounding, unlike the IPW estimator. In scenarios where the estimators were approximately unbiased, CI widths were comparable regardless of the point and variance estimators used.

**Table 1:** Average empirical bias and 95% confidence interval (CI) coverage and width for the IPW estimator and the regression estimators using sampling weights (Reg Wts) and offsets (Reg Offset). Variances were estimated using the Taylor Series (TS) and bootstrap (Boot) variance estimators, and for IPW the bootstrap estimator accounting for propensity score estimation (Boot+PS) is also shown. Results are given for both the marginal subtype conditional OR for subtype 1 (*OR*_1_) and subtype 2 (*OR*_2_). Scenarios shown are for 1% sampling proportion of first phase observations into second phase. Empirical bias values larger than 0.1 and 95% CI coverage values less than 0.90 are underlined.

| Scenario | Method | Variance<br>Est Method | Bias |  | Coverage |  | CI Width |  |
| --- | --- | --- | --- | --- | --- | --- | --- | --- |
| | | | $OR_1$ | $OR_2$ | $OR_1$ | $OR_2$ | $OR_1$ | $OR_2$ |
| $OR_1 = 1.00, OR_2 = 1.00,$<br>No confounding | IPW | Boot+PS | 0.03 | 0.01 | 0.97 | 0.94 | 0.96 | 0.43 |
|  |  | Boot |  |  | 0.97 | 0.95 | 0.97 | 0.43 |
|  |  | TS |  |  | 0.97 | 0.94 | 0.95 | 0.43 |
|  | Reg Wts | Boot | 0.03 | 0.01 | 0.97 | 0.95 | 0.97 | 0.43 |
|  |  | TS |  |  | 0.97 | 0.95 | 0.95 | 0.43 |
|  | Reg Offset | Boot | 0.03 | 0.01 | 0.96 | 0.95 | 0.95 | 0.42 |
|  |  | TS |  |  | 0.96 | 0.95 | 0.93 | 0.42 |
| $OR_1 = 0.86, OR_2 = 0.18,$<br>Confounding<br>with interactions | IPW | Boot+PS | 0.00 | 0.01 | 0.95 | 0.96 | 0.35 | 0.30 |
|  |  | Boot |  |  | 0.97 | 0.97 | 0.40 | 0.30 |
|  |  | TS |  |  | 0.97 | 0.96 | 0.40 | 0.28 |
|  | Reg Wts | Boot | <u>0.27</u> | 0.01 | <u>0.32</u> | 0.97 | 0.49 | 0.30 |
|  |  | TS |  |  | <u>0.32</u> | 0.95 | 0.49 | 0.29 |
|  | Reg Offset | Boot | <u>0.19</u> | 0.03 | <u>0.56</u> | 0.96 | 0.46 | 0.33 |
|  |  | TS |  |  | <u>0.58</u> | 0.95 | 0.46 | 0.31 |

### 3.2 CBCS Analysis

The CBCS data were used to evaluate differences in the subtype-conditional exposure effect estimates for BMI, breastfeeding, oral contraceptive use, and parity. Figure 1 displays the estimated ORs and 95% CIs for each exposure of interest using each method outlined in Section 2. The variances for the estimated subtype-conditional ORs from each method were estimated using both the Taylor series and nonparametric bootstrap estimators, as well as the bootstrap estimator accounting for propensity score estimation for the IPW estimator. For each of the exposures considered, the CIs for the regression estimator with offsets were narrower than those of the other estimators. Variance estimates were similar for each subtype-conditional OR estimator. Slight differences in subtype-conditional OR estimates and their estimated variances were observed across methods, with the most pronounced differences occurring for parity. Frequency distributions of breast cancer subtype within each exposure are also given in Figure 1. Note for parity that the cell counts for the nulliparous group were small, particularly for the *ER–/p53+* subtype, contributing to the larger width of the CI for the corresponding OR.

**Figure 1:**
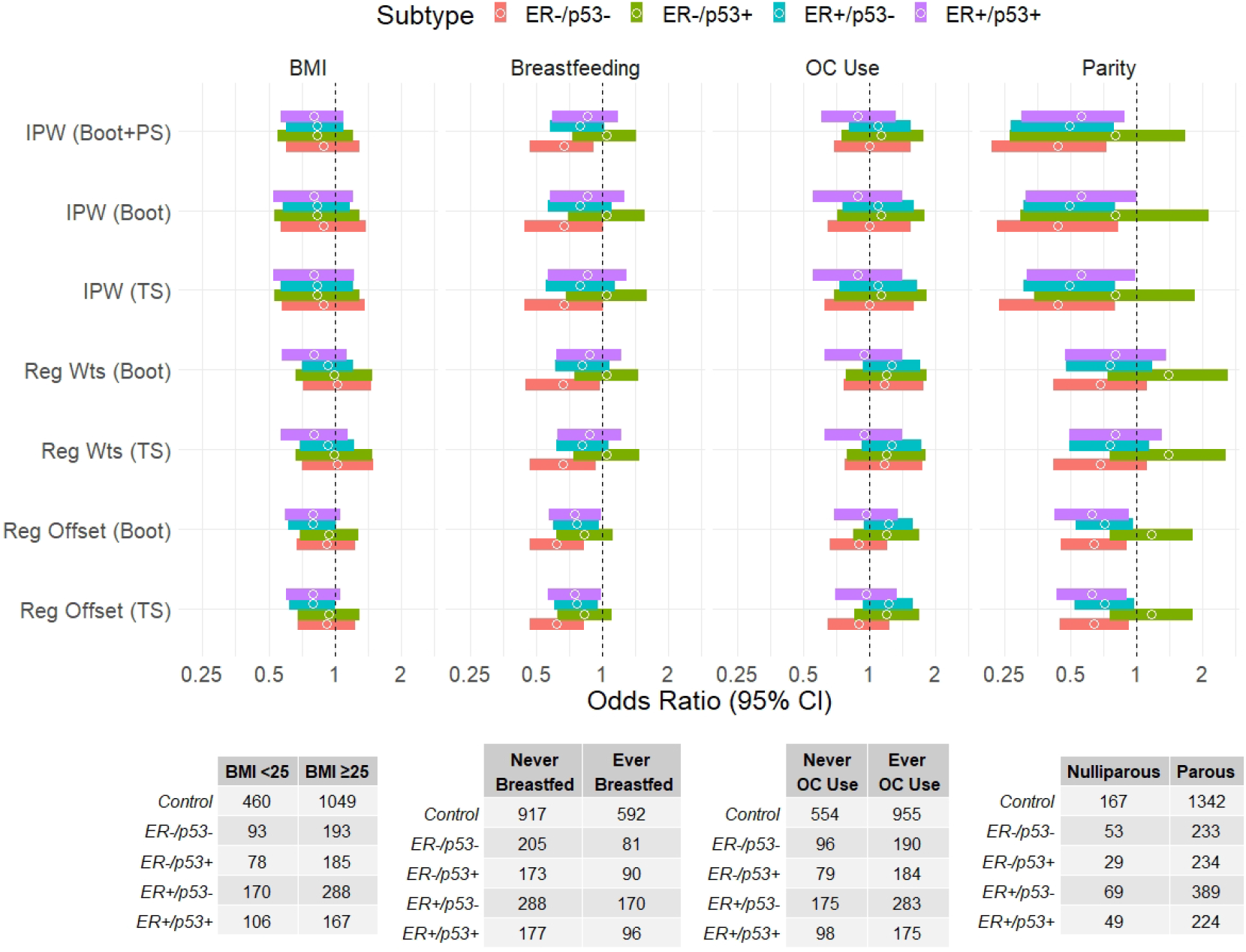
Subtype-conditional odds ratio estimates and 95% confidence intervals (CIs) for BMI (referent: *<* 25), breastfeeding (referent: never), oral contraceptive (OC) use (referent: never), and parity (referent: nulliparous). Odds ratios and CIs estimated using each of the methods presented in Section 2. Contingency tables of the exposure-outcome frequency distributions are displayed beneath their corresponding column of the plot.

The distribution of the weights estimated in the IPW approach were examined for each exposure and trimming of weights was considered. In general, the mean of the estimated weights should be close to one and there should not be extreme weights for any individual if the propensity score model is correctly specified (Cole and Hernán, 2008). The estimated weights corresponding to the BMI, breastfeeding, and oral contraceptive use analyses each met these criteria. For the parity analysis, the mean of the estimated weights was 1.31 and there were some individuals with extreme weights (range: 0.3, 80.4). The estimated weights were trimmed to the 99^*th*^ percentile (i.e., weights greater than 10.1 were set to this value) in an effort to reduce possible bias and inflated variance of the estimated exposure effect. After trimming, the estimated ORs remained similar and their corresponding CIs were narrower; the results of the parity analysis with trimmed weights are displayed in Figure 1.

Table 2 gives p-values for the test of no heterogeneity of exposure effect across breast cancer subtypes, for each of BMI, breastfeeding, oral contraceptive use, and parity. Specifically, each p-value corresponds to the F-test of *H*_0_ : *OR*_−−_ = *OR*_−+_ = *OR*_+−_ = *OR*_++_ for the given exposure, where *OR*_−+_ represents the subtype-conditional OR for subtype *ER–/p53+*, and ORs for the other subtypes are defined analogously. Recall that for regression the ORs are covariate-conditional, whereas for IPW the ORs have a marginal interpretation. Overall, none of the p-values for the test of heterogeneity were less than 0.05 but there were some differences between methods. For instance, for parity the *F*-test p-values corresponding to IPW were several times larger than p-values corresponding to the regression approaches. Within each modeling approach, variance estimation methods performed similarly for each exposure. Finally, note that while Table 2 provides p-values across a number of exposures without subsequent adjustment, multiple testing should generally be taken into account for such analyses involving multiple comparisons.

**Table 2:** P-values for the test of no heterogeneity across subtypes for each of BMI, breastfeeding, oral contraceptive (OC) use, and parity.

| Method | Var. Est. | BMI | Breastfeeding | OC Use | Parity |
| --- | --- | --- | --- | --- | --- |
| IPW | Boot+PS | 0.98 | 0.26 | 0.75 | 0.69 |
|  | Boot | 0.97 | 0.25 | 0.65 | 0.67 |
|  | TS | 0.97 | 0.25 | 0.69 | 0.65 |
| Reg Wts | Boot | 0.77 | 0.29 | 0.68 | 0.27 |
|  | TS | 0.75 | 0.26 | 0.71 | 0.21 |
| Reg Offset | Boot | 0.71 | 0.57 | 0.90 | 0.09 |
|  | TS | 0.70 | 0.47 | 0.87 | 0.07 |

## 4 Discussion

The proposed IPW method can be used to simultaneously account for confounding and two-phase ODS when the outcome is multinomial. On the other hand, regression with sampling weights or offsets will not yield unbiased estimates of marginal subtype-conditional ORs in general in the presence of confounding. This was demonstrated in the simulation studies, where the regression approaches at times showed substantial bias of the OR estimator and poor CI coverage. For CBCS, similar results were observed across methods for BMI, breastfeeding, and oral contraceptive use, but substantial differences were observed across methods for parity. All methods considered here modeled subtypes jointly. Alternatively outcome subtypes may be modeled separately, although this approach doesn’t allow for estimation of the covariances between the subtype-conditional ORs, precluding inference comparing subtype-conditional ORs. Fitting the multinomial models described in Section 2 facilitates between subtype comparisons, allowing for estimation of the covariance matrix and therefore testing null hypotheses involving multiple subtype-conditional ORs. Further, applying IPW in this setting enables inference about effect heterogeneity, yielding approximately unbiased estimates of marginal subtype-conditional ORs and valid CIs in the presence of confounding and biased sampling.

The CBCS analysis presented in Section 3 has some limitations. Firstly, the sample size of CBCS is moderate in the sense that certain breast cancer subtypes are rare and therefore the number of individuals with these subtypes is small; for instance, there are only 29 nulliparous individuals with *ER–/p53+* breast cancer in Phase I and II of CBCS. Additionally, in the analyses of Section 3 the assumption that the variables *L* were sufficient for confounding adjustment may not hold, and this could result in invalid inference using each of the methods considered. For example, inclusion of parity in the set *L* for the analysis of oral contraceptive use is debatable because parity could be on the causal path between oral contraceptive use and breast cancer. The CBCS data was recorded at a single time point for each person, but the true relationship between parity and oral contraceptive use is likely cyclic and best described with longitudinal data. In this case, part of the effect of parity in the CBCS analysis may be mediated by adjusting for oral contraceptive use. Alternatively, not adjusting for oral contraceptive use may result in residual confounding of the effect of parity on breast cancer subtype. Due to this complicated relationship, longitudinal data is recommended to answer the questions about parity posed in this research. In the CBCS analysis it was assumed that the Medicare and DMV records data constituted a random sample from the population of women in central NC. If the individuals not included in these records were systematically different than those included in the records, the sample would be biased in a manner that is not adjusted for by the methods presented. In turn, the OR estimators from each of the methods may be biased estimates of the effects of the exposures considered in the population.

There are several possible areas of related research which could be undertaken. For instance, future work could explore improving efficiency by utilizing data from individuals not selected into the second phase. In particular, the simple and enriched doubly robust estimators and the locally efficient estimator presented in Wang et al. (2009) for two-phase ODS may be adapted to the multinomial outcome setting. Finally, further investigation of the potential consequences of extreme sampling weights and violations (or near-violations) of positivity is desirable.

## Data Availability

All data produced in the present study are available upon reasonable request to the authors.

## Appendix A Details of Methods Presented in the Main Text

### A.1 Propensity Score Model

The approach described in Wang et al. (2009) is used to estimate the propensity score. Assume the logistic regression model logit *e*_1_(*L*; *α*) = *h*(*L*; *α*) where *h*(*L*; *α*) is some function linear in *α*, e.g., *h* {*L*; *α* = (*α*_0_, *α*_1_)} = *α*_0_ + *α*_1_*L*. The corresponding vector of score functions for this model can be expressed *∂h*(*L*; *α*)*/∂α* {*A* − *e*_1_(*L*; *α*)}. In order to account for the second stage of sampling, each individual is weighted using weights *S/g*(*Y, L*_1_, *A*). Fitting this model by maximizing the weighted likelihood with the given weights yields the estimator 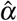, where 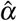is the solution to

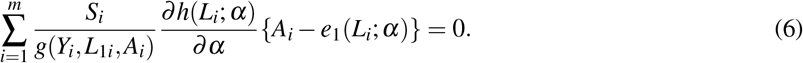

Note that there is complete data for *n* of the *m* individuals from the first-phase sample (those with *S*_*i*_ = 1), and the remaining *m* − *n* do not contribute to the sum in (6). The estimating equations vector (6) is unbiased, i.e.,

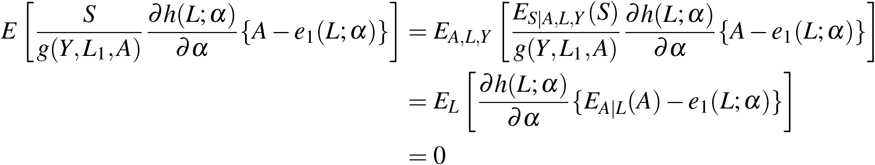

implying 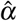 is a consistent and asymptotically normal estimator of *α*.

### A.2 Inverse Probability Weighting

Consider first the case where *K* = 1, i.e., *Y* is binary. Wang et al. (2009) propose the following inverse probability weighting approach to causal effect estimation in the context of two-phase outcome-dependent sampling studies. For estimation of the counterfactual means *µ*_*a*_ = *E*[*Y*^*a*^], *a* = 0, 1, note that the following is a mean zero estimating function:

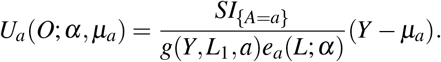

This suggests estimating *µ*_*a*_ by the solution to

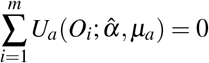

which yields the Hajek-type estimator

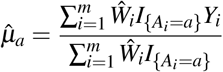

that is consistent for *µ*_*a*_, since the corresponding vector of estimating equations for 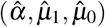is unbiased.

Now consider the multinomial case where *K >* 1. Let *π* = (*π*^1^, *π*^0^) where 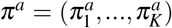 for *a* = 1, …, *K* and *a* = 0, 1, the 2*K* estimating functions for the counterfactual probabilities can be written

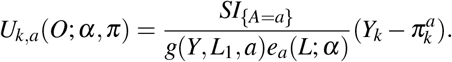

Let *W*_*i*_ = *S*_*i*_ {*A*_*i*_*/e*_1_(*L*_*i*_; *α*) + (1 − *A*_*i*_)*/e*_0_(*L*_*i*_; *α*) }*/g*(*Y*_*i*_, *L*_1*i*_, *A*_*i*_). The vector of estimating functions *Ψ*(*O*_*i*_; *α, π*) for the IPW estimator is obtained by stacking the score functions for the propensity score model with the estimating functions for the counterfactual probabilities:

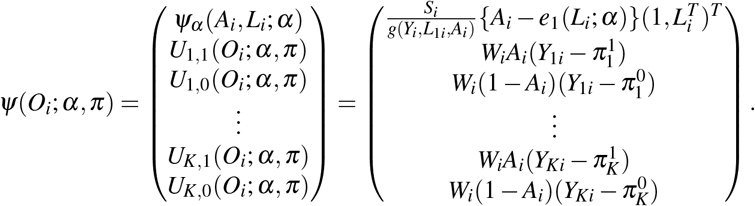

The estimator 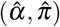 that solves ∑_*i*_ *Ψ*(*O*_*i*_; *α, π*) = 0 is consistent for (*α, π*) and asymptotically normal because the estimating equations vector is unbiased. This was demonstrated in Section A.1 above for *Ψ*_*α*_ (*A*_*i*_, *L*_*i*_; *α*), and for *U*_*k*,1_(*O*_*i*_; *α, π*) we have

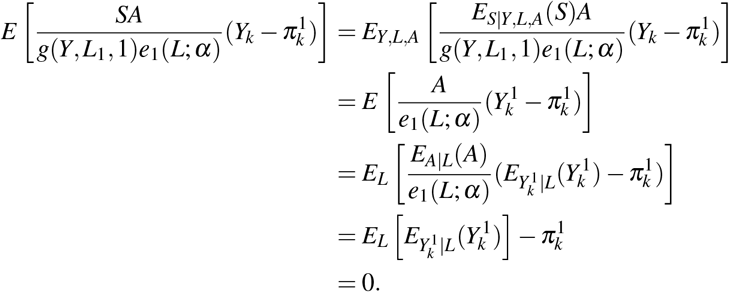

The proof for *U*_*k*,0_(*O*_*i*_; *α, π*) is analogous.

In terms of the parameters from the MSM in Section 2.1 of the main text and the propensity score model from Section A.1, we have the following vector of estimating functions

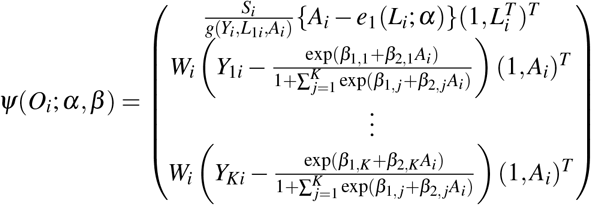

where 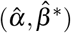is a solution to the unbiased estimating equations vector ∑_*i*_ *Ψ*(*O*_*i*_; *α, β*) = 0. Thus 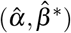is consistent for (*α, β*) and asymptotically normal.

### A.3 Taylor Series Variance Estimator

The Taylor Series (TS) variance estimator is commonly used for complex survey data, where sampling may be done by clusters (primary sampling units) or within strata defined by certain variables. The following is an explanation of the structure of the TS variance estimator used in SAS Software, and how its form can be simplified to the TS variance estimator referenced in this manuscript.

Let *π*_*hi jk*_ represent the probability of developing outcome subtype *k* where *h* = 1, …, *H* is the stratum index, *i* = 1, …, *n*_*h*_ is the cluster index within stratum *h*, and *j* = 1, …, *m*_*hi*_ is the unit index within cluster *i* of stratum *h*. The multinomial model can be written

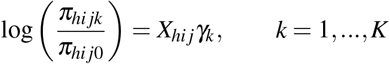

where *γ*_*k*_ = (*γ*_*k*1_, …, *γ*_*kp*_)^*T*^ and 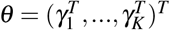The TS variance estimator used in the SAS Software Version 9.4 *SURVEYLOGISTIC* procedure can be expressed as 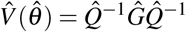where

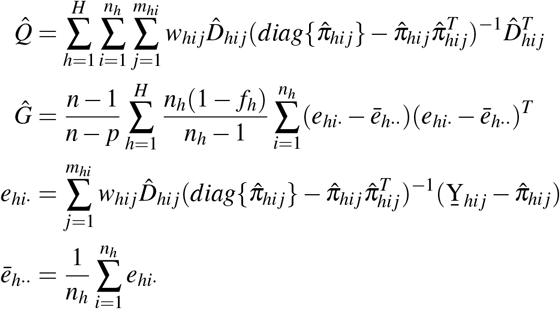

and 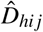 is the matrix of partial derivatives of the function *g*(*X*_*hi j*_, *θ*) = *π*_*hi j*_ with respect to *θ* evaluated at 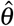, *w*_*hi j*_ is a scalar weight, *f*_*h*_ is the sampling rate for stratum *h*, and 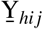 is the column vector of length *K* whose elements are indicator variables for the categories 1, …, *K* of variable Y. Let 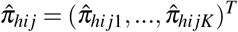where 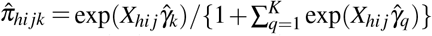and 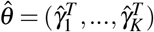is the vector of MLEs from fitting the multinomial model at the beginning of this section.

The TS variance estimator 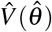is commonly used for complex survey data, and the formula can be simplified for the purposes of this manuscript. In particular, for *n* i.i.d. individuals let *H* = 1 so there is only one stratum, let *f*_*h*_ = 0, and let each individual *i* represent their own cluster such that *n*_*h*_ = *n* and *m*_*hi*_ = 1. Under these conditions, each component may be expressed without the subscripts *h* and *j*. Noting that 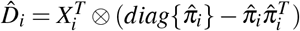), we have 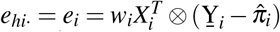 which represents the score function vector of length × *K* evaluated at 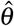 *p*. Then 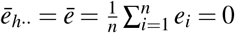 and

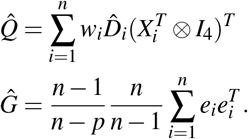

Now note the summand in 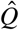 is equal to the negative derivative of the score function vector, evaluated at 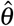. The summand in 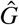 is equal to the outer product of the score function vector, evaluated at 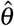. Thus 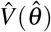 is proportional to the Huber-White (HW) robust sandwich variance estimator 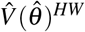. Specifically, 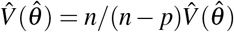. If fitting the multinomial models presented in Section 2 of the main text using the *SURVEYLOGISTIC* procedure in SAS, the option *VADJUST=NONE* may be used in the *MODEL* statement to suppress the adjustment factor (*n* − 1)*/*(*n* − *p*), yielding the unadjusted TS variance estimator 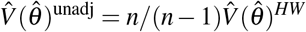.

### A.4 Regression with Sampling Weights

Fitting model (2.5) from the main text by maximizing the weighted likelihood with weights equal to *S*_*i*_*/g*(*Y*_*i*_, *L*_1*i*_, *A*_*i*_) yields the estimator 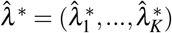, where 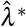is the solution to

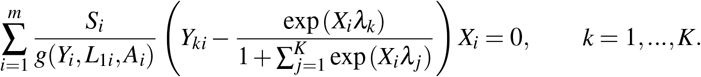

This estimating equations vector is unbiased, i.e.,

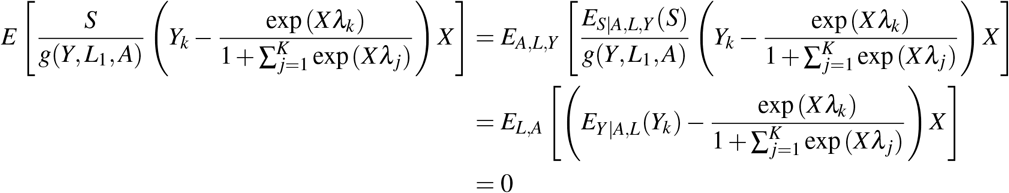

implying 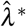 is a consistent and asymptotically normal estimator of *λ*. Note *E*_*Y*|*A,L*_(*Y*_*k*_) = *E*_*Y*|*X*_ (*Y*_*k*_) = *P*(*Y* = *k*|*X*) and 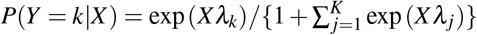 from model (2.3) in the main text.

### A.5 Regression with Offsets

Consider first the case where *Y* is binary and *X* is defined as in the main text. We wish to estimate the parameters in the following model,

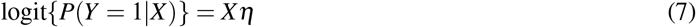

but we have a biased sample and cannot fit this model directly. Weinberg and Wacholder (1990) propose the following model that can be fit with the observed data

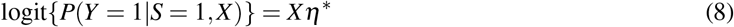

where *Xη*^∗^ = *Xη* + *Xω* and *Xω* is the offset term and *ω* is a known column vector. Given this relationship, the maximum likelihood estimator for *η* can be expressed as 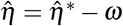, where 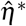 is the maximum likelihood estimator for *η*^∗^. In other words, model A.3 can be fit to obtain consistent and asymptotically normal estimators of the parameters of model A.2.

The model with offsets above yields the following mean zero vector of score functions:

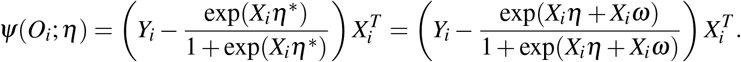

Weinberg and Wacholder (1990) also note that since the offset is dependent only on the data *X* and the known constant *ω*, the covariance matrix is consistently estimated with no correction required.

Now let *Y* be multinomial taking values *k* = 0, …, *K, K >* 1. Assume selection was performed with respect to the dichotomized outcome; modification of this approach may be required for study designs in which selection is performed with respect to individual subtypes.

Assume the model (2.5) from the main text, where 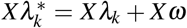and *Xω* represents the known vector of individual offset terms defined in equation (2.4) of the main text. Note that

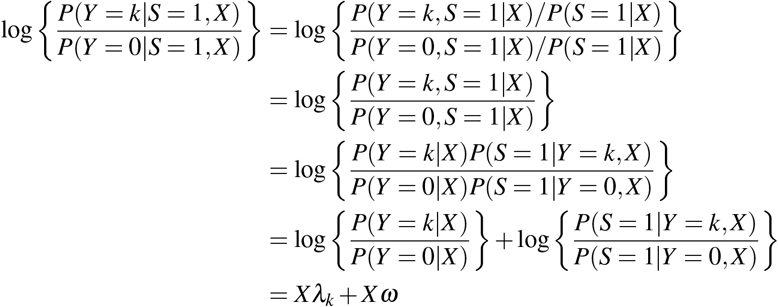

for all *k* = 1, …, *K*. The above equation holds because *P*(*S* = 1 *Y* = *k, X*) = *P*(*S* = 1 *Y* = *k, L*_1_ = *l*_1_, *A* = *a*), since it is assumed that *S* ⊥ *L*_2_ ⊥ *Y, L*_1_, *A*. Applying the same logic as above for the binary case, fitting model (2.5) by maximizing the likelihood including the offset term results in consistent and asymptotically normal estimators of the *λ*_*k*_ from model (2.3).

## Appendix B Additional Simulation Study Results

Table 3 below displays simulation study results for the same scenarios given in Table 1 of the main text, but with 10% sampling of first-phase observations into the second phase.

**Table 3:** Average empirical bias and 95% confidence interval (CI) coverage and width for the IPW estimator and the regression estimators using sampling weights (Reg Wts) and offsets (Reg Offset). Variances were estimated using the Taylor Series (TS) and bootstrap (Boot) variance estimators, and for IPW the bootstrap estimator accounting for propensity score estimation (Boot+PS) is also shown. Results are given for both the marginal subtype conditional OR for subtype 1 (*OR*_1_) and subtype 2 (*OR*_2_). Scenarios shown are for 10% sampling proportion of first-phase observations into the second phase. Empirical bias values larger than 0.1 and 95% CI coverage values less than 0.90 are underlined.

| Scenario | Method | Variance<br>Est Method | Bias |  | Coverage |  | CI Width |  |
| --- | --- | --- | --- | --- | --- | --- | --- | --- |
| | | | $OR_1$ | $OR_2$ | $OR_1$ | $OR_2$ | $OR_1$ | $OR_2$ |
| $OR_1 = 1.05, OR_2 = 1.06$,<br>No confounding | IPW | Boot+PS | 0.00 | -0.01 | 0.96 | 0.97 | 0.98 | 0.44 |
|  |  | Boot |  |  | 0.96 | 0.97 | 0.99 | 0.44 |
|  |  | TS |  |  | 0.96 | 0.97 | 0.98 | 0.44 |
|  | Reg Wts | Boot | 0.00 | -0.02 | 0.96 | 0.97 | 1.00 | 0.44 |
|  |  | TS |  |  | 0.96 | 0.97 | 0.98 | 0.44 |
|  | Reg Offset | Boot | -0.01 | -0.02 | 0.96 | 0.96 | 0.97 | 0.43 |
|  |  | TS |  |  | 0.96 | 0.96 | 0.95 | 0.43 |
| $OR_1 = 0.86, OR_2 = 0.17$,<br>Confounding<br>with interactions | IPW | Boot+PS | -0.04 | 0.01 | 0.93 | 0.96 | 0.33 | 0.33 |
|  |  | Boot |  |  | 0.96 | 0.97 | 0.37 | 0.36 |
|  |  | TS |  |  | 0.97 | 0.95 | 0.37 | 0.27 |
|  | Reg Wts | Boot | <u>0.21</u> | 0.01 | <u>0.52</u> | 0.97 | 0.46 | 0.29 |
|  |  | TS |  |  | <u>0.51</u> | 0.96 | 0.46 | 0.27 |
|  | Reg Offset | Boot | <u>0.13</u> | 0.03 | <u>0.77</u> | 0.96 | 0.43 | 0.31 |
|  |  | TS |  |  | <u>0.79</u> | 0.95 | 0.43 | 0.29 |

## Appendix C SAS Workflow

This section presents details and SAS code for analyzing a simulated data set similar to those used in the second simulation study presented in the main text. In particular, the scenario demonstrated here corresponds to the simulation study with a 1% sampling proportion, where there is confounding with interactions and *OR*_1_ = 0.86, *OR*_2_ = 0.18.

### C.1 Simulated Data Set

#### C.1.1 First-Phase Data

The first-phase data set contains *m* = 200, 000 individuals that were simulated according to the following specifications. Recall that *A* represents the observed exposure, and *L*_1_, *L*_2_ were the covariates observed in the first and second phases, respectively. Their distributions for each individual were specified as follows:

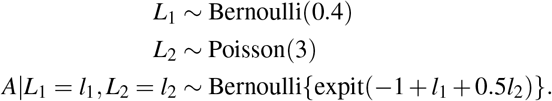

The counterfactual disease subtype under exposure *a, Y*^*a*^, has three levels and was generated according to the following distribution:

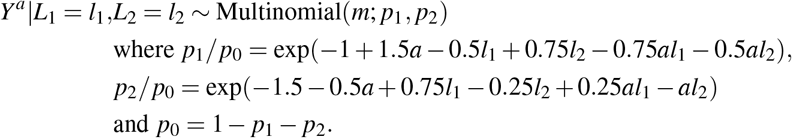

Note that *p*_1_ is the probability of developing disease subtype 1 given the covariates *L*_1_, *L*_2_ under exposure *a*, and *p*_0_, *p*_2_ have analogous interpretations for the reference subtype (i.e., subtype 0) and subtype 2, respectively. Then the observed outcome for each individual was *Y* = *AY* ^1^ +(1 − *A*)*Y* ^0^. Unlike in the CBCS data, the outcome *Y* here has 3 levels and *L*_1_, *L*_2_ each represent only one variable.

#### C.1.2 Defining Sampling Weights and Selecting Second-Phase Data

The second-phase data set contained *n* = 2, 000 individuals selected from the first-phase data. The sampling weights were defined using probabilities of selection based on the joint distribution of *Y, L*_1_ in the first-phase population, to mirror the CBCS data. In particular, let *d* _*jk*_ = *m P*(*L*_1_ = *j, I*(*Y >* 0) = *k*) for *j, k* ∈ {0, 1} where the *d* _*jk*_ sum to the size of the first-phase population, i.e.,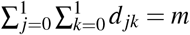.

The second-phase selection probabilities were defined for *j* = 0, 1 as *P*(*S* = 1 |*L*_1_ = *j,Y* = 0) = 0.25*n/d*_*j*0_ for those with outcome subtype 0 and *P*(*S* = 1 |*L*_1_ = *j,Y* = *k*) = 0.25*n/d*_*j*1_ for those with subtypes 1 or 2; these probabilities were defined as such in order to sample an approximately equal number of individuals from the four groups. As in the main text, the selection weight for each individual was then the inverse of their selection probability.

For the simulation studies described in the main text, the sampling weights were used to draw 500 second-phase samples of size 2,000 from the first-phase data. One such data set was analyzed in the following sections for the purpose of this workflow. This data set was named dat and had one row per individual in the second-phase sample (2, 000 rows total), and had variables *L*1, *L*2, *A,Y*, and *wt*_*sel* (selection weights).

### C.2 Model Fitting and Hypothesis Testing

#### C.2.1 IPW

After a second-phase data set was selected, the propensity score model was fit and the IP weights were computed as shown below. Using IP weights, the model in equation (2.2) of the main text was fit to estimate the parameters of the marginal structural model described in Section 2.1.

~~~
*Fit propensity score model ;
proc logistic data=dat ;
  class A(ref=“0”) L1 ;
  model A = L1 L2 ;
  weight wt_sel ;
  output out=ps predicted=ps_pred ;
run ;
*Construct IP weights ;
data dat_ps ;
  set ps ;
  if A=0 then ipw = 1 / (1 - ps_pred) ;
  else if A=1 then ipw = 1 / ps_pred ;
  combo_wt = wt_sel*ipw ;
run ;
*Estimate parameters of MSM ;
proc surveylogistic data=dat_ps varmethod=taylor ;
   class Y A(ref=“0”) / param=ref ;
   model Y(ref=“0”) = A / link=glogit covb ;
   weight combo_wt ;
   NoHet: test A1_1=A1_2 ;
run ;
~~~

The test statement performed the F-test of *H*_0_ described in Section 2.1. The covb option specified in the model statement output the estimated covariance matrix, and was used to identify the labels SAS gave the model parameters; these were needed for specification of the null hypothesis in the test statement.

The variance estimation method shown in the code above corresponds to the Taylor Series method described in the main text, and this is the default method for this procedure. To use the bootstrap variance estimator instead, use `varmethod=bootstrap`. Note that this is the bootstrap method that does not account for propensity score estimation. The bootstrap variance estimator that accounts for propensity score estimation is not currently available in PROC SURVEYLOGISTIC, but can be called using the *%boot* macro (can be downloaded here: https://github.com/friendly/SAS-macros/blob/master/jackboot.sas) as shown below. Likewise, the F-test using this variance estimator can be performed manually using PROC IML.

~~~
%include “YourDirectoryPath\jackboot.sas” ; /* file containing %boot macro */
*Bootstrap estimator accounting for propensity score estimation ;
%boot(data=dat,
   samples=250,
   random=12345,
   chart=0,
   stat=Int_1 Int_2 Est_1 Est_2,
   alpha=0.05,
   print=1) ;
*Manually perform F-test using output data sets from %boot ;
%let df_den = 1999 ; /* Denominator degrees of freedom */
%let df_num = 1 ; /* Numerator degrees of freedom */
data statout ; /* Get parameter estimates in correct order */
  set bootstat (where=(name in (“Int_1”, “Int_2”)))
    bootstat (where=(name in (“Est_1”, “Est_2”))) ;
   keep value ;
run ;
ods output PredCov=V_pbin ; /* Output covariance matrix for parameter estimates */
proc calis data=bootdist pcorr ;
   mstruct var= Int_1 Int_2 Est_1 Est_2 ;
run ;
proc iml ;
   reset log print ;
   use V_pbin ;
   read all into V ; /* Covariance matrix */
   close V_pbin ;
   L = {0 0 1 -1} ; /* Define contrast matrix */
   use statout ;
   read all into beta ; /* Estimated parameter vector */
   close statout ;
   /* Compute F-statistic */
Fstat = ((&df_den.-&df_num.+1)/(&df_den.*&df_num.)) *
         (L * beta)’ * inv(L * V * L’) * (L * beta) ;
  create outF var {Fstat} ;
  append ;
  close outF ;
quit ;
data outp ; /* Get corresponding p-value */
  set outF ;
  p = 1 - CDF(‘F’, Fstat, &df_num., &df_den.) ;
run ;
~~~

#### C.2.2 Regression with Sampling Weights or Offsets

The sampling weights have been described above in terms of known conditional selection probabilities, and the offset term was defined using these probabilities as well. In particular, following equation (2.4) from the main text, the offset term was log {*P*(*S* = 1 | *L*_1_ = *j,Y* = *k*)*/P*(*S* = 1 *L*_1_ = *j,Y* = 0) }for *j* = 0, 1. Then the regression model in equation (2.5) was fit in two ways, as demonstrated below, to estimate the parameters in equation (2.3).

~~~
*Fit Regression model with sampling weights ;
proc surveylogistic data=dat varmethod=taylor ;
    class Y A(ref=“0”) L1 / param=ref ;
    model Y(ref=“0”) = A L1 L2 / link=glogit covb ;
    weight wt_sel ;
    NoHet: test A1_1=A1_2 ;
run ;
*Fit Regression model with offset term ;
proc surveylogistic data=dat varmethod=taylor ;
   class Y A(ref=“0”) L1 / param=ref ;
   model Y(ref=“0”) = A L1 L2 / link=glogit offset=off covb ;
   NoHet: test A1_1=A1_2 ;
run ;
~~~

As mentioned for IPW, the variance estimation specified here corresponds to the Taylor Series method. The F-test of no heterogeneity within strata defined by *L* was invoked here with the test statement.

## References

Ahearn, T. U., Zhang, H., Michailidou, K., Milne, R. L., Bolla, M. K., Dennis, J., Dunning, A. M., Lush, M., Wang, Q., Andrulis, I. L., et al. (2022). Common variants in breast cancer risk loci predispose to distinct tumor subtypes. Breast Cancer Research, 24(1):1–13.

Benefield, H. C., Zabor, E. C., Shan, Y., Allott, E. H., Begg, C. B., and Troester, M. A. (2019). Evidence for Etiologic Subtypes of Breast Cancer in the Carolina Breast Cancer Study. Cancer Epidemiology, Biomarkers & Prevention, 28(11):1784–1791.

Binder, D. A. (1983). On the variances of asymptotically normal estimators from complex surveys. International Statistical Review, 51(3):279–292.

Chatterjee, N. (2004). A two-stage regression model for epidemiological studies with multivariate disease classification data. Journal of the American Statistical Association, 99(465):127–138.

Cole, S. R. and Hernán, M. A. (2008). Constructing inverse probability weights for marginal structural models. American Journal of Epidemiology, 168(6):656–664.

Greenland, S., Robins, J. M., and Pearl, J. (1999). Confounding and collapsibility in causal inference. Statistical Science, 14(1):29–46.

Mashreghi, Z., Haziza, D., and Léger, C. (2016). A survey of bootstrap methods in finite population sampling. Statistics Surveys, 10:1–52.

Newman, B., Moorman, P. G., Millikan, R., Qaqish, B. F., Geradts, J., Aldrich, T. E., and Liu, E. T. (1995). The Carolina Breast Cancer Study: integrating population-based epidemiology and molecular biology. Breast Cancer Research and Treatment, 35(1):51–60.

Neyman, J. (1938). Contribution to the Theory of Sampling Human Populations. Journal of the American Statistical Association, 33(201):101–116.

Richardson, D. B., Kinlaw, A. C., Keil, A. P., Naimi, A. I., Kaufman, J. S., and Cole, S. R. (2018). Inverse Probability Weights for the Analysis of Polytomous Outcomes. American Journal of Epidemiology, 187(5):1125–1127.

Rosner, B., Glynn, R. J., Tamimi, R. M., Chen, W. Y., Colditz, G. A., Willett, W. C., and Hankinson, S. E. (2013). Breast cancer risk prediction with heterogeneous risk profiles according to breast cancer tumor markers. American Journal of Epidemiology, 178(2):296–308.

SAS Institute Inc. (2017). User’s Guide The SURVEYLOGISTIC Procedure. In SAS/STAT 14.3 User’s Guide, chapter 114, pages 9328–9428. SAS Institute Inc., Cary, NC.

Sul, J. H., Martin, L. S., and Eskin, E. (2018). Population structure in genetic studies: Confounding factors and mixed models. PLoS Genetics, 14(12):e1007309.

Wang, M., Kuchiba, A., and Ogino, S. (2015). A meta-regression method for studying etiological heterogeneity across disease subtypes classified by multiple biomarkers. American Journal of Epidemiology, 182(3):263–270.

Wang, W., Scharfstein, D., Tan, Z., and MacKenzie, E. J. (2009). Causal inference in outcome-dependent two-phase sampling designs. Journal of the Royal Statistical Society. Series B: Statistical Methodology, 71(5):947–969.

Weinberg, C. R. and Wacholder, S. (1990). The Design and Analysis of Case-Control Studies with Biased Sampling. Biometrics, 46(4):963–975.

Zabor, E. C. and Begg, C. B. (2017). A comparison of statistical methods for the study of etiologic heterogeneity. Statistics in Medicine, 36(25):4050–4060.

Zhang, H., Ahearn, T. U., Lecarpentier, J., Barnes, D., Beesley, J., Jiang, X., O’Mara, T. A., Qi, G., Zhao, N., Bolla, M. K., et al. (2020). Genome-wide association study identifies 32 novel breast cancer susceptibility loci from overall and subtype-specific analyses. Nature Genetics, 52(6):572–581.

Zhang, H., Zhao, N., Ahearn, T. U., Wheeler, W., García-Closas, M., and Chatterjee, N. (2021). A mixed-model approach for powerful testing of genetic associations with cancer risk incorporating tumor characteristics. Biostatistics, 22(4):772–788.

